# Expanding the Phenotypic spectrum of *NAA10*-related neurodevelopmental syndrome and *NAA15*-related neurodevelopmental syndrome

**DOI:** 10.1101/2022.08.22.22279061

**Authors:** Gholson J. Lyon, Marall Vedaie, Travis Besheim, Agnes Park, Elaine Marchi, Leah Gottlieb, Tzung-Chien Hsieh, Hannah Klinkhammer, Katherine Sandomirsky, Hanyin Cheng, Lois J. Starr, Isabelle Preddy, Marcellus Tseng, Quan Li, Yu Hu, Kai Wang, Ana Carvalho, Francisco Martinez, Alfonso Caro-Llopis, Maureen Gavin, Karen Amble, Peter Krawitz, Ronen Marmorstein, Ellen Herr-Israel

## Abstract

Amino-terminal (Nt-) acetylation (NTA) is a common protein modification, affecting 80% of cytosolic proteins in humans. The human essential gene, *NAA10,* encodes for the enzyme NAA10, which is the catalytic subunit in the N-terminal acetyltransferase A (NatA) complex, also including the accessory protein, NAA15. The full spectrum of human genetic variation in this pathway is currently unknown. Here we reveal the genetic landscape of variation in *NAA10* and *NAA15* in humans. Through a genotype-first approach, one clinician interviewed the parents of 56 individuals with *NAA10* variants and 19 individuals with *NAA15* variants, which were added to all known cases (N=106 for *NAA10* and N=66 for *NAA15*). Although there is clinical overlap between the two syndromes, functional assessment demonstrates that the overall level of functioning for the probands with *NAA10* variants is significantly lower than the probands with *NAA15* variants. The phenotypic spectrum includes variable levels of intellectual disability, delayed milestones, autism spectrum disorder, craniofacial dysmorphology, cardiac anomalies, seizures, and visual abnormalities (including cortical visual impairment and microphthalmia). One female with the p.Arg83Cys variant and one female with an *NAA15* frameshift variant both have microphthalmia. The frameshift variants located toward the C-terminal end of *NAA10* have much less impact on overall functioning, whereas the females with the p.Arg83Cys missense in NAA10 have substantial impairment. The overall data are consistent with a phenotypic spectrum for these alleles, involving multiple organ systems, thus revealing the widespread effect of alterations of the NTA pathway in humans.

## Introduction

Targeting 40% of the human proteome, NatA, the major N-terminal acetyltransferase (NAT) complex, acetylates Ser-, Ala-, Gly-, Thr-, Val-, and Cys- N-termini following cleavage of the initiator methionine ^1, 2^. The canonical human NatA consists of two main subunits, the catalytic subunit N-α-acetyltransferase 10 (NAA10) (Ard1) and the auxiliary subunit NAA15 (Nat1) and engages with a regulatory subunit, HYPK ^3–5^. NAA15 function has been linked to cell survival, tumor progression, and retinal development ^6, 7^. N-terminal (Nt-) acetylation (NTA) is one of the most common protein modifications, occurring co- and post-translationally ^8^. Approximately 80% of cytosolic proteins are N-terminally acetylated in humans and ∼50% in yeast ^1^, while NTA is less common in prokaryotes and archaea ^8^.

NTA is catalyzed by a set of enzymes, the NATs, which transfer an acetyl group from acetyl coenzyme A (Ac-CoA) to the free α-amino group of a protein’s N-terminus. To date, eight distinct NATs (NatA – NatH) have been identified in metazoan (NatA-F and NatH) and plant (NatG) species that are classified based on different conserved subunit compositions and substrate specificities. NTA has been implicated in steering protein folding, stability or degradation, subcellular targeting, and complex formation ^9^. Particularly, Naa10-catalyzed N-terminal acetylation has been reported to be essential for development in many species and although NatA is not essential in *S. cerevisiae*, depletion of Naa10 or Naa15 has strong effects, including slow growth and decreased survival when exposed to various stresses. In addition, it has been recently shown that mice have a compensating enzyme Naa12, which prevents embryonic lethality in the *Naa10* knockouts ^10^, but a similar gene has not been found in humans. Furthermore, *NAA10* was also identified in screens for essential genes in human cell lines ^11, 12^.

Ogden syndrome (OS) was first reported in 2011, and it was given this name due to the location of the first affected family residing in Ogden, Utah, USA ^13, 14^. In that first family, five males were born to four different mothers and all the males died in early infancy, with a range of cardiac and other defects, including lethal cardiac arrhythmias. The underlying genetic defect was characterized: this variant involved a single missense change coding for Ser37Pro in the X-linked gene, *NAA10*, which was found in a second independent family in California, USA with three males that had also died during infancy. The identical variant was recently reported in a third family ^15^. There is a *S. cerevisiae* model for the Naa10 Ser37Pro mutant, in which that variant impairs NatA complex formation and leads to a reduction in both NatA catalytic activity and functionality ^16, 17^. Furthermore, OS patient-derived cells have impaired *in vivo* NTA of a few NatA substrates ^18^.

Since the initial discovery of OS in 2011, multiple groups have reported additional variants either in *NAA10* in both males and females^19–27^ or in the heterodimeric protein partner encoded by *NAA15* ^20, 28, 29^ (although reference limitations at the journal do not allow us to cite all relevant papers, so we are only citing a few here, but see **Supplementary Discussion**). Given that NAA10 and NAA15 are part of the NatA complex, it is very likely that there might be shared phenotypes in individuals with pathogenic variants in either of them. As such, the goal of the present study was for one clinician-scientist (G.J.L.) to meet many families with *NAA10* and *NAA15* variants directly and prospectively to uncover patterns that might have been missed by the prior retrospective reviews of medical records and/or summaries provided by the referring clinicians. The prospective collection of phenotyping information presented here, through a set of direct interviews with the families, has produced a better overall understanding of the range of organ systems affected and the overall natural history of these conditions. In addition, a subset of these families underwent functional assessment of the probands, with one psychologist (E.I.) administering the Vineland-3 to all patients coming to the Jervis clinic for developmental assessments.

## Materials and Methods

### Clinical features methodology

New families were either self-referred, referred by their clinicians, or referred by social media Facebook group parent managers to the corresponding author (GJL). Consent forms were provided to the families for a research study collecting medical information and, for those families who provided written consent, an in-person assessment or videoconference assessment was performed with each family by G.J.L. The in-person assessments included nine visits by *NAA10* families from September 2019 to March 2020, but these visits stopped with the onset of the COVID19 pandemic in March 2020. During these nine visits, the research participants were examined by a medical geneticist, a neurologist, a psychiatrist, and an optometrist. Phenotypic information was also obtained from clinical records with varying data available, ranging from a list of the key clinical features to detailed history and examination findings. A separate written informed consent was obtained for publication of photographs.

A summary template was formulated and filled in for each research participant, where this template was first completed by a research coordinator (E.M.) and then reviewed by a second person for accuracy. These summaries are available from the corresponding author, as MedRxiv does allow posting of such information, and these summaries will be submitted as Supplementary Information for eventual publication in a peer-reviewed journal. The phenotype information from all published and unpublished cases was uploaded to the Human Disease Gene website series ^30^ with secondary data quality control completed by a second person as well, to ensure accuracy and completeness. Human Phenotype Ontology (HPO) terms were selected in the Human Disease Gene website and the percentages represent the collation of groups of multiple terms falling under one feature archetype. For example, “Motor delay” included all probands listed as having “Gait disturbance”, “Fine Motor Delay”, “Gross Motor Delay”, “Difficulty Standing” or “Inability to stand” entered into their summary. While the intent of this method was to create a more simplified and comprehensive gestalt of the OS patient, the natural limitation to this method is the lack of nuance in distinguishing minute differences in presentation from one patient to another. Another limitation may be the overlap of symptoms falling between different feature archetypes, such as “Poor eye contact” being heavily associated with (but not exclusive to) “autistic behavior”, the latter intended to reflect more stereotyped and repetitive behavior. Future research may wish to parse out the fine distinctions between these behaviors for greater clarity. Both pertinent positive and negative features were noted, with phenotypes classified as “pertinent negatives” only if the provided clinical information explicitly stated that a phenotype was denied by the parents, was noted to be absent during physical examinations, or not reported during diagnostic procedures. Clinical features were only reported as pertinent negatives if this was explicitly mentioned in the records, otherwise, these were classified as “unknown” or “not available”. The relative prevalence of each phenotype was calculated by dividing the number of individuals positive for the phenotype by the sample size. Percentiles for head circumference, weight and height were calculated using CDC growth charts. On the website, for *NAA10*, the protein variants were numbered with three significant digits, thus allowing sorting in order from 1 to 235 amino acids; for example, p.Arg83Cys. For *NAA15*, the cDNA and protein variants were numbered with four digits, also to allow sorting.

### Variant identification and Bioinformatics Methodology

Variants were identified primarily using exome sequencing through clinical diagnostic testing. The sequencing kits and technology varied based on the different companies involved, and the variants of interest were highlighted and verified in the collected clinical diagnostic test reports.

For the annotation of ClinVar variants, the combined annotation dependent depletion (CADD) scores, MPC scores, Polyphen-2, SIFT scores, M-CAP_score and GERP++_RS were retrieved from the dbNSFP4.3a database. Consensus scoring for “pathogenic” was assigned when CADD-scores were above 23, MPC scores were>=2, GERP++_RS score were >=2, and SIFT, Polyphen and M-CAP all provided supportive evidence for pathogenicity for these variants. The variant was assigned by consensus as benign when more than 3 above approaches provided evidence of benign or tolerance. We also classified variants according to the ACMG Guidelines and the Association for Molecular Pathology (ACMG/AMP) framework ^31^. For gnomAD frequency, the population maximum frequency is set at 4.066e-06 in Annovar, so a period in that column (**Supplementary Table 4**) indicates that the allele is less frequent than this.

### Molecular Analysis of NAA10 and NAA15 Mutations

Molecular effects of the mutations in NAA10 and NAA15 subunits were discussed based upon the human NatA crystal structure (PDB: 6C9M) and the human NatA/HYPK crystal structure (PDB: 6C95), with the heterodimeric NatA crystal structure being used as a representative model for the prepared figures. The structures were visually inspected using PyMol, which was also used in conjunction with Adobe Illustrator CC to prepare **Figure 4** and **Figure 5**. The discussion was also informed by numerous biochemical and biophysical studies of NAA10 and NAA15 mutant proteins ^13, 16, 18–21, 23, 32, 33^.

### Patient clustering analysis description

Phenotypes (manually curated HPO terms and identifiers) of patients were extracted using Perl and taken as input of clustering analysis. Resnik similarity score ^34^ was calculated between any two HPO terms using the Python module ssmpy ^35^. Given each pair of patients, the similarity score is formalized as:

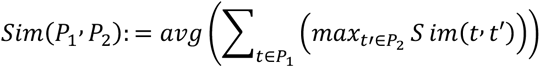

where *P*_1_ and *P*_2_ represent patient 1 and 2, and *t* and *t*’ represent HPO terms in patient 1 and 2. Similarity score matrix between patients were taken as input of software R. Patient-patient difference were calculated based on 1/Resnik similarity score. Hierarchical clustering analysis was performed using hclust function in R based on patient-patient difference. Hierarchical clustering results was visualized as dendrogram in R. Patients from NAA10 and NAA15 were highlighted using different colors. In addition to tree structure, we visualized the clustering results using multidimensional scaling (MDS) based on patient-patient difference matrix. Specifically, we assume *x_i_* = (*x_i_*_1_, …, *x_in_*)in n-dimensional space for each patient *i* and estimate coordinate matrix *X* = (*x_i_*, …, *x_N_*) for all patients by minimizing strain function:

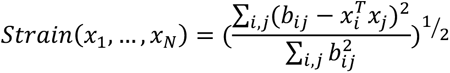

where *b_ij_* are element of the double centralized patient-patient difference matrix *B* = 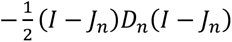, where *I* is identity matrix, *J_n_* is all-one matrix and *D_n_* is patient-patient distance square matrix. Then, to minimize the strain function above, singular value decomposition is applied to matrix *B*. Patient-coordinate matrix 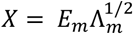 where *E_m_*. is the matrix of *m* eigenvectors of *B* and Λ_m_. is the diagonal of *m* eigenvalue of *B*. To visualize the patient clustering results, we reduce dimension of *X* to two by selecting top two largest eigenvalue of *E_m_* All process is implemented in R.

### Facial Gestalt Methodology

To validate whether two cohorts share a similar facial phenotype, we conducted a statistical analysis in the clinical face phenotype space of GestaltMatcher ^36, 37^. In this analysis, we analyze 78 NAA10 patients and 33 NAA15 patients, including the patients reported in this work and the previously published patients in the GestaltMatcher Database (GMDB; https://db.gestaltmatcher.org). The information on all patients performed in this analysis is shown in **Supplementary Table 6**. The mean pairwise cosine distance between patients from the two cohorts is examined to measure the similarity. We sampled control distributions of mean pairwise distances between two cohorts stemming a) from the same syndrome and b) from two different syndromes (shown in blue (a) and red (b) in **Supplementary Figure 8**. Cohorts were sampled from patients in the GMDB that were not included in the training of GestaltMatcher. Then, a ROC analysis was conducted to derive a threshold to decide whether two cohorts stem from the same or different syndromes. The control distributions and resulting threshold were tuned via a five-fold cross-validation resulting in a final threshold of *c* = 0.9176. To test the similarity of the two cohorts *C*_1_ and *C*_2_, their mean pairwise cosine distance *d*(*C*_1_, *C*_2_) is calculated and compared to *c*. Additionally, we sample two subcohorts from *C*_1_ and *C*_2_ respectively and calculate their mean pairwise cosine distance for 100 times. If at least 50% of those 100 subsampled comparisons fall above the threshold *c*, it is considered as evidence for the two cohorts stemming from different syndromes. The approach was tested on the validation fold not used in the derivation of *c* comprising 66 syndromes. The method correctly detected two cohorts stemming from the same syndrome in 80.30% of the comparisons and correctly identified two cohorts stemming from different syndromes in 91.98% of the comparisons.

### Rating scales

The Vineland-3 Comprehensive Interview Form was administered to the parents by one psychologist (E.I.), and provides norm-referenced scores at three levels: subdomains, domains, and the overall Adaptive Behavior Composite (ABC). Adaptive behavior subdomains make up the most fine-grained score level. The primary norm-referenced scores for the subdomains are v-scale scores, which have a mean of 15 and standard deviation (SD) of 3. The v-scale score for each subdomain is included in the narrative interpretation. Standard scores have a mean of 100 and SD of 15. Confidence intervals reflect the effects of measurement error and provide, for each standard score, a range within which the proband’s true standard score falls with a certain probability or confidence. The confidence level chosen for the report is the 90% confidence interval. A percentile rank is the percentage of individuals in the age group who scored the same or lower than the proband. For example, a percentile rank of 41 indicates that the examinee scored higher than (or the same as) 41% of the age-matched norm sample.

## Results

An in-person assessment was performed on nine families with *NAA10* variants from October 2019 – March 2020 and an additional 26 families with *NAA10* variants were interviewed by videoconferencing from April 2020 until September 2020 due to the COVID19 pandemic. All families seen by September 2020 were scheduled for cognitive evaluations by one qualified psychologist (E.I.). For families with *NAA15* variants, none of the families were seen in-person prior to pandemic onset and ten families were seen via videoconferencing from April 2020 until September 2020, also with referral for cognitive evaluation. After this time, from October 2020 until July 2021, an additional 20 families with *NAA10* variants and 9 families with *NAA15* variants were seen via videoconferencing, and cognitive evaluations are being conducted on an ongoing basis for these families. One other family with a recently deceased child with a *NAA10* variant provided extensive medical records and consent to publish but was unable to meet for videoconferencing. All summary data and percentages for phenotype data for these and other published (and some unpublished) *NAA10* and *NAA15* cases was curated at the Human Disease Gene website series ^30^, under the gene names *NAA10* and *NAA15*, at the following links using the “Graph and Chart” tab: https://humandiseasegenes.nl/naa10/ and https://humandiseasegenes.nl/naa15/

The data were downloaded from the website on May 2, 2022; these results are shown in **Supplementary Table 1** (an excel file, with tabs for *NAA10* and *NAA15*). The website series is meant to be an open-source venue for such data ^30^. World maps showing the location of the cases for *NAA10* and *NAA15* are included in the public-facing portion of the websites.

### NAA10 and NAA15 variants

The numbering of individuals and the variants are shown in **Figure 1**, **Supplementary Table 2** for *NAA10* and in **Supplementary Table 3** for *NAA15*. The overwhelming majority of individuals seen were female, where one missense variant (c.247C>T, p.Arg83Cys) in *NAA10* occurs more frequently and is most recurrent in females, totaling to 25 females seen in this study. However, consistent with this X-linked disease being more severe in males, one male (Individual 9) with the p.Arg83Cys missense in NAA10 died at 11 months of age. Other variants were recurrent, but at a much lower frequency, including p.Tyr43Ser, p.Ile72Tyr, p.Ala87Ser, p.Phe128Leu, and p.Met147Thr. In this cohort of individuals, novel variants include: p.His16Pro, p.Tyr31Cys, p.Ala104Asp, p.Arg116Trp, p.His120Pro, p.Leu121Val, p.Ser123Pro, p.Phe128Ser (same position as p.Phe128Leu), p.Thr152Argfs*6, and p.Glu181Alafs*67. New information was gathered on every individual reported herein, while a shorter summary was published previously describing some of these individuals with *NAA10* variants (specifically individuals 8, 11, 13-21, 35-38, 41, 47, and 51) ^20^.

**Figure 1.**
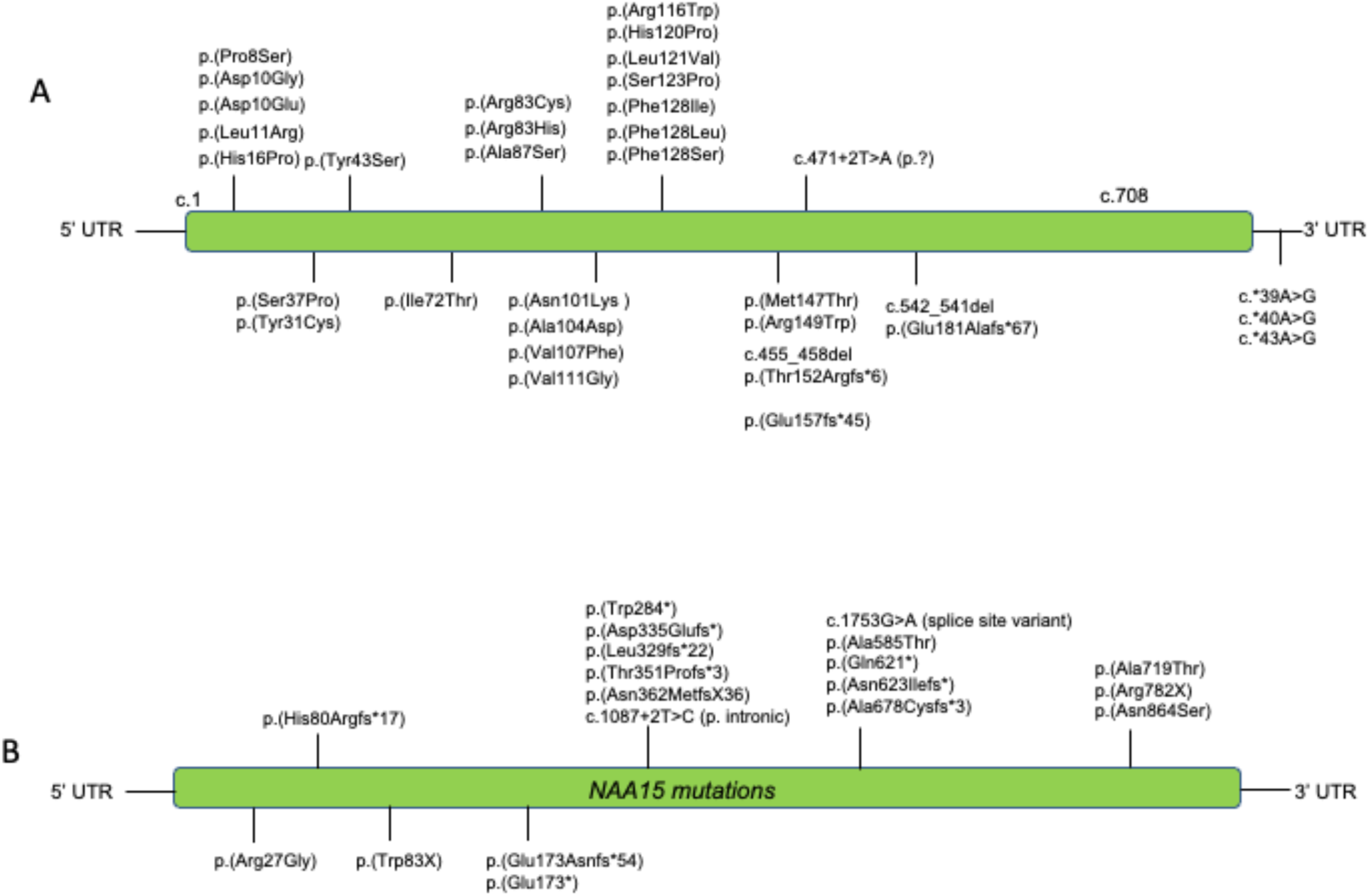
Pathogenic variants in *NAA10* and *NAA15* reported herein. Abbreviations: UTR: untranslated region.

One child (Individual 1) was found to have two *de novo* variants on the same sequencing reads: [c.22C>T;30C>G] p.[Pro8Ser;Asp10Glu]. The c.022C>T variant affects the first base of exon 2 with possible interference on the splicing of messenger RNA. Clinical studies of the messenger RNA expression in the patient confirmed that mRNA from this allele was not present (**Supplementary Figure 1**). The clinical presentation for this child is very similar to many other individuals with other *NAA10* variants, further supporting a haploinsufficiency model in which loss of function of one allele seems to mirror the effects of missense variants in other individuals.

Most of the *NAA10* variants are *de novo*, with the exception of p.Tyr43Ser in a previously reported family ^38^, p.Ile72Thr in two families (one of which, Individual 8, was previously reported ^20, 23^ and Individual 54 with frameshift variant p.Thr152Argfs*6, with this same p.Thr152Argfs*6 variant being reported in two other cases ^20, 39^.

For *NAA15*, further information about the variants is supplied in **Supplementary Table 3**. Two of these individuals (4 and 19) were published previously and referred to as Individuals 14 and 21 in that paper ^28^. Almost all variants are *de novo*, with the exception of a splice site variant with an unknown inheritance pattern.

As of April 2022, there are 58 putative missense or in-frame deletion, substitution or insertion variants in *NAA10* and 64 putative missense or in-frame deletion, substitution or insertion variants in *NAA15* submitted to ClinVar, with many of these listed as variants of uncertain significance (**Supplementary Table 4**). The bioinformatic analyses described in the Methods section reports consensus classification for each variant, with an upgrade to pathogenic for several of these. However, many of these entries come from clinical laboratories, with very little uploaded phenotype information, and future work will involve an attempt to contact and interview these other families. Of course, all of the information in ClinVar is de-identified, so this requires contacting the clinical testing laboratories and/or other sources of the data, and then getting referred to the clinicians who are involved in these cases. For now, it is worth at least cataloging what is currently in ClinVar, along with the new bioinformatic annotations.

### Clinical Features

Although some variants will be discussed in detail below, most individuals with variants involving either *NAA10* or *NAA15* have variable degrees of neurodevelopmental disabilities, including impaired motor abilities (HP:0001270), ID (HP:0001249), impaired verbal abilities (HP:0000750) and autism spectrum disorder (ASD) (HP:0000729) (**Table 1**). These data include previously published cases with variants in *NAA10*, so the overall numbers are greater than just the families that were videoconferenced. Due to having only one X-chromosome, the males with variants in *NAA10* are usually much more severely affected with cardiac issues to the point where some of these individuals died in infancy. However, it is notable that one male, Individual 7 with p.Ile72Thr has no apparent cardiac issues and his level of functioning (as assessed by Vineland-3) was better than some of the females. This is in contrast to a different child (Individual 8) with the exact same variant who experienced a sudden cardiac death around 5 years of age and had prior development of a medulloblastoma ^20^. In the cohort, the only other individual developing any type of cancer was Individual 2 with a p.His16Pro missense in NAA10, with leukemia cured by chemotherapy, although a case report of a child with the same exact variant did not report any leukemia or other cancers ^19^. As such, this very low level of cancer with just one case of medulloblastoma and one case of leukemia in this current cohort could be explained simply by coincidence or other unknown genetic causes not yet identified, but, given the literature regarding the involvement of *NAA10* perhaps in cancer development ^40^, this likely warrants further investigation. Overall, individuals with *NAA10* variants more often have much more impaired motor function, including fine motor difficulties, abnormality of movement, motor delay, and hypotonia, in comparison to individuals with *NAA15* variants.

**Table 1.** Phenotype feature frequency in Individual with *NAA10* variants.

| System | Feature | Prevalence (% positive/(negative + unknown)) |
| --- | --- | --- |
| Neurologic | •Intellectual disability<br>•Delayed speech<br>•Motor delay<br>•Developmental delay<br>•Hypotonia<br>•Seizures<br>•Corpus callosum hypoplasia<br>•Microcephaly<br>•Periventricular leukomalacia | 92/95 (96.8%)<br>91/95 (95.8%)<br>89/95 (93.7%)<br>77/95 (81.1%)<br>57/95 (60.0%)<br>34/95 (35.8%)<br>27/95 (28.4%)<br>25/85 (26.3%)<br>17/95 (17.9%) |
| Psychiatric | •Harming behaviors (self or others)<br>•Attention deficits<br>•Eye contact deficits<br>•Autistic behavior<br>•Compulsive behavior<br>•Impulsive behavior | 37/95 (38.9%)<br>31/85 (32.6%)<br>28/95 (29.5%)<br>26/85 (27.4%)<br>16/95 (16.8%)<br>11/95 (11.6%) |
| Cardiovascular | •Prolonged QT interval<br>•Atrial septal defects<br>•Ventricular septal defects<br>•Hypertrophic cardiomyopathy<br>•Arrhythmias<br>•Murmurs<br>•Cardiac arrest | 17/95 (17.9%)<br>17/95 (17.9%)<br>13/95 (13.7%)<br>9/95 (9.5%)<br>9/95 (9.5%)<br>8/95 (8.4%)<br>6/95 (6.3%) |
| Respiratory | •Neonatal respiratory distress<br>•Pneumonia & other recurrent upper respiratory infections<br>•Apnea<br>•Pulmonary hypertension<br>•Bronchiolitis | 14/95 (14.7%)<br>14/95 (14.7%)<br>5/95 (5.3%)<br>5/95 (5.3%)<br>4/95 (4.2%) |
| Gastrointestinal | •Feeding difficulties in infancy<br>-Gastrostomy tube feeding<br>-Nasogastric tube feeding<br>•Feeding difficulties after infancy<br>•Dysphagia<br>•Gastroesophageal reflux<br>•Vomiting<br>•Bowel incontinence<br>•Constipation<br>•Diarrhea | 57/95 (60.0%)<br>12/95 (12.6%)<br>10/95 (10.5%)<br>40/95 (42.1%)<br>27/95 (28.4%)<br>21/95 (22.1%)<br>19/95 (20.0%)<br>16/95 (16.8%)<br>9/95 (9.5%)<br>7/95 (7.4%) |
| Eye abnormalities | •Abnormal eyelashes<br>•Thick eyebrows<br>•Abnormal palpebral fissure<br>•Strabismus<br>•Hypertelorism | 42/95 (44.2%)<br>35/95 (36.8%)<br>22/95 (23.1%)<br>18/95 (18.9%)<br>17/95 (17.9%) |
| Visual abnormalities | •Astigmatism<br>•Myopia<br>•Cortical visual impairment | 22/95 (23.1%)<br>17/95 (17.9%)<br>13/95 (13.7%) |
| Facial features | •Ear abnormalities<br>-Low-set<br>-Other outer ear abnormalities<br>-Middle and internal ear abnormalities<br>•Nose abnormalities<br>-Abnormal nasal bridge<br>-Abnormal nasal tip<br>-Ala or nares abnormalities<br>•Mouth abnormalities<br>-Abnormal philtrum, vermillion<br>-Abnormal palate<br>-Teeth and gingival abnormalities<br>•Chin abnormalities | 50/95 (52.6%)<br>32/95 (33.7%)<br>35/95 (36.8%)<br>6/95 (6.3%)<br>64/95 (67.4%)<br>39/95 (41.1%)<br>48/95 (50.5%)<br>12/95 (12.6%)<br>69/95 (72.6%)<br>59/95 (62.1%)<br>29/95 (30.5%)<br>29/95 (30.5%)<br>9/95 (9.5%) |
| Metabolic & Endocrine | •Sleep disturbance<br>-Sleep apnea<br>•Abnormality of temperature regulation<br>•Slender build<br>•Hirsutism<br>•Precocious puberty | 28/95 (29.5%)<br>4/95 (4.2%)<br>28/95 (29.5%)<br>25/95 (26.3%)<br>14/95 (14.7%)<br>6/95 (6.3%) |

Various levels of ID are reported in almost all study subjects with available data, including mild, moderate or severe ID, and learning difficulties with or without behavioral issues. The severity of the intellectual disability, when known, for *NAA10* and *NAA15* is shown in **Figure 2** and shows that *NAA10* cases are usually much more severe than *NAA15* cases, which is once again consistent with X-linked inheritance for *NAA10*, along with the fact that NAA10 is the acetyltransferase enzyme, whereas NAA15 is the dimeric binding partner that helps to localize the NatA complex to the ribosome.

**Figure 2.**
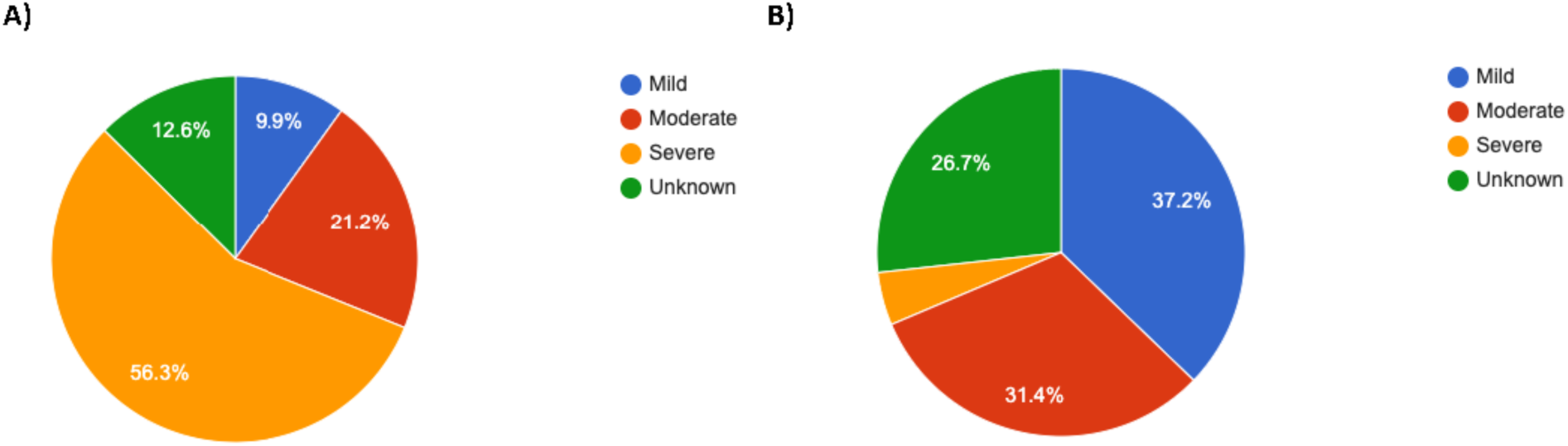
Severity of Intellectual Disability. A) Individuals with *NAA10* variants (n=95); B) Individuals with *NAA15* variants (n=57).

The two male individuals with frameshift variants in *NAA10* (Individuals 55 and 56) are generally much higher functioning than the other individuals with missense variants. For example, Individual 56 was reported to be cognitively better than individuals his age (for example, he can count from 1 to 10 and knows almost all the letters of the alphabet and is only 29 months of age). Although it was noted that he is a couple of months behind in gross motor skills (he still cannot run or jump even though he tries. He does attend a typical nursery and the staff report that he is intelligent, and “he always wants to learn something else”. His teacher says that while she “has to explain something to individuals in his age 10 times to get them to do it, he already does it the second time”. These frameshift variants occur toward the C-terminus of the protein, with Individual 56 being the most C-terminal with Glu181Alafs*67 in *NAA10*. Therefore, it is likely that some functional N-terminal acetyltransferase enzymatic activity remains.

Most individuals with *NAA10* variants have verbal issues including complete absence of speech, delayed language development, required use of sign language, or other speech difficulties related to articulation, which some parents report as possibly being connected to low muscle tone in the pharyngeal muscles. Some research participants also present with either ASD and/or other behavioral challenges. The birth weight was low (≤ 5th percentile) in a few individuals and it was noted that some individuals failed to grow, causing their weights to fall below the 5th percentile over time. The percentages for some clinical features are listed in **Table 1** and can be found on the Human Disease Gene Website series.

The most notable new information relates primarily to the development and maturation of these individuals, as it has become apparent that there are several hurdles to overcome during development, and that failure to overcome each hurdle significantly impacts the trajectory of development. First and foremost, these individuals must overcome the neonatal lethality of possible cardiac arrythmias and cardiovascular or respiratory complications. This seems to be more common in males, but one female (Individual 3) with a p.Tyr31Cys missense in NAA10 died at 6.5 months with such issues after spending some time in the neonatal intensive care unit (NICU) (see **Supplementary Case** information), which also occurred with male infants in the past, including the male Individual 9 reported herein. The second hurdle involves feeding difficulties in infancy including swallowing food and liquids, as some individuals have severe hypotonia, and ultimately require feeding tube insertions ^41^. The third hurdle involves remaining seizure-free, as it seems that the cognitive development of the individuals is markedly delayed if seizures begin and are not well-controlled. Lastly, the fourth hurdle involves learning to walk, as some of the individuals remain wheelchair-bound, due to a lack of coordination and/or an inability to stay balanced when walking, which seems to also be related to low muscle tone. As such, the clinical presentation can be quite variable, as someone who requires tube-feeding, is wheelchair bound, and is suffering from seizures will likely have a far different cognitive outcome than someone who does not experience these setbacks.

### Facial features

Although it was previously noted that “a recognizable, regular pattern of dysmorphic facial features was not appreciated amongst the cases” ^36, 37^, this is debatable now that one clinician has met and examined these individuals via videoconferencing, as there are commonly seen and sometimes recognizable traits in individuals with *NAA10* variants, including thicker eyebrows, long eyelashes, upturned nose, and broad nasal bridge (**Table 1**, **Supplementary Figure 2-4**). While the facial features in the males and females can also be quite variable, some, including an upturned nose, along with bushy eyebrows and long eyelashes, are quite common. Eye abnormalities were found in almost all individuals, which sometimes included large, prominent eyes and large down-slanting palpebral fissures. Other possible features include delayed closing of anterior and posterior fontanels, presence of a broad and/or prominent forehead, and sparse anterior scalp hair. The most common ear findings were low-set ears and large ears, with a broad range of hearing impairment and abnormalities of the outer ear. Nose features included broad nasal bridge, prominent or broad nasal tip, and anteverted or flared nares. Lastly, mouth abnormalities were present in a majority of the individuals with a high palate being the main feature, but there was a range of abnormalities in the vermillion and philtrum, as well as teeth abnormalities with particular issues with eruption and disorganized growth. There were several cases with short chins, protruding upper lip, and microretrognathia.

Facial feature abnormalities also vary in many individuals with *NAA15* variants (**Supplementary Figure 5**). The most common are abnormalities of the eyebrows (horizontal, thick, or long), long eyelashes, hypertelorism, prominent epicanthal folds, abnormalities of the palpebral fissure, amblyopia, astigmatism, and/or strabismus. Mouth features were also common with abnormalities of the philtrum (long, short, broad, or smooth), teeth, upper lip vermillion, chin, and/or presence of a high palate. Abnormalities of the nose included prominent nasal tip or abnormal nasal bridge. Ear features were least common but presented mostly as outer ear abnormalities or low-set ears.

To investigate further the similarity between the two cohorts based on facial gestalt, we conducted a facial analysis using the GestaltMatcher approach^41^, using all available facial photographs for *NAA10* and *NAA15* patients, including previously published cases (**Supplementary Table 6**). We first utilized tSNE ^42^ to visualize the distribution of *NAA10* and *NAA15* patients in the two-dimensional space. In **Supplementary Figure 7**, we can see that *NAA10* and *NAA15* patients do not separate clearly. Moreover, comparing the cohorts *NAA10* and NAA15 in the clinical face phenotype space resulted in a mean pairwise cosine distance of *d*(*NAA*10, *NAA*15) = 0.9146 between two patients. The mean pairwise distance falls below the threshold *c* = 0.9176 defined to decide whether two cohorts stem from the same syndrome (mean pairwise cosine distance smaller than *c*) or from two different syndromes (mean pairwise cosine distance greater than *c*). A subsampling approach yielded that 67% of sampled subcohorts showed similar phenotypes. Overall, our analysis indicates an overlap in the facial gestalt of NAA10 patients and NAA15 patients (**Supplementary Figure 8**).

### Cardiovascular features

The majority of females and even some males have no obvious cardiac issues. However, several of the males had structural anomalies of their hearts, including ventricular septal defect, atrial septal defect, and pulmonary artery stenosis. For those who died, arrhythmias at the time of death included torsade de pointes, premature ventricular contraction (PVC), premature atrial contraction (PAC), supraventricular tachycardia (SVtach), and ventricular tachycardia (Vtach). Death can result from cardiogenic shock following arrhythmia, which was noted in some affected individuals.

Baseline echocardiography is recommended as septal defects have been identified in several cases. Medical (pharmaceutical) management has been employed to slow the progression of congestive heart failure (CHF) when manifesting in some individuals. Electrocardiography (EKG) is also obtained at the time of diagnosis, as fatal dysrhythmias can develop with time. In several cases, the initial EKG has been normal, with evolution of arrhythmias during later periods of life. It may be necessary to evaluate on an annual basis and be vigilant for signs of and symptoms of dysrhythmia/CHF. When present, the arrhythmias have proven difficult to manage, though the full spectrum of anti-arrhythmics has not been tested. Individuals have responded to episodic cardioversion/defibrillation, but long-term treatment has not yet been determined. Pacemakers and implantable defibrillators have been used in a few cases, including in one published family ^38^.

In addition to gross anatomical defects that subsequently resulted in arrythmias, EKG analysis of OS patients has demonstrated a large proportion of patients demonstrating an elongated QT interval congruent with previously published literature ^38^. Our data shows an equal proportion of patients with elongated QT intervals as patients with atrial septal defects in patients with variants in *NAA10* (**Table 1**). These seem to be well-controlled with pharmacotherapy in these patients and caught routinely with Holter monitor assessment.

### Respiratory features

The vast majority of probands do not have any major respiratory issues, but there are a few isolated cases of interstitial lung disease and/or respiratory distress (**Table 1**). For example, Individual 3 with p.Tyr31Cys missense in NAA10 died in the first year of life with local interstitial lung disease, neonatal respiratory distress, respiratory failure requiring assisted ventilation, lymphangiectasia, and pulmonary fibrosis. This included repeated apnea episodes requiring CPR after one such episode. Radiological CT imaging of the thorax was performed at 6 weeks of age and reported “ground glass changes throughout both lungs”. The etiology remained uncertain and a lung biopsy reported mild thickening of the interstitium without significant interstitial inflammatory infiltrate, but expansion of the inter-lobular septae with lymphangiectasia and fibrosis. Focal interstitial glycogenosis was confirmed with PAS staining. The Individual 9, a male with p.Arg83Cys missense in NAA10, died around one year of age, also with respiratory arrest, neonatal respiratory distress, respiratory failure requiring assisted ventilation, tachypnea, and interstitial lung disease. Of course, the original OS males also had respiratory distress and frequent pneumonias ^13^, seen also in a third family ^15^, but these other individuals without primary cardiac arrhythmias seem to demonstrate that this respiratory distress can be primarily caused by interstitial lung disease, rather than being entirely secondary to cardiovascular complications.

As mentioned above, most individuals did not have any notable respiratory issues, but a recognizable proportion of our cohort did demonstrate a pattern of recurrent respiratory infections similar to the original OS males ^13^, with varying severity and frequency. A larger proportion of patients with *NAA10* variants had respiratory infections as opposed to individuals with *NAA15* variants, and this pattern is replicated with the other respiratory features. A few patients also had neonatal respiratory distress, pulmonary hypertension and/or apnea (such as Individuals 12, 38, 45, 47 and 56). Given the relative lack of respiratory distress in most cases, many clinical tests, such as CT scanning and/or lung biopsy were not clinically indicated, so these data are simply not available. As such, it is not known if these individuals never had any interstitial lung disease or if they perhaps had a mild case which subsequently resolved over time. In this regard, it was written that one female with p.His16Pro in *NAA10* needed postnatally to “have an oxygen mask applied at night because of oxygen desaturations”, and “because of symptoms resembling interstitial lung disease (chronic tachy-dyspnea, recurrent pneumonia and bronchitis), a lung biopsy was performed at the age of 3 years without revealing any specific findings” ^40^. As such, it is possible that some of these individuals might have interstitial lung disease neonatally, but that in many cases, this might resolve over time.

### Gastrointestinal features

The gastrointestinal pathology associated with OS, in order from most to least prevalent, includes feeding difficulties in infancy, dysphagia, GERD/silent reflux, vomiting, constipation, diarrhea, bowel incontinence, and presence of eosinophils on esophageal endoscopy. The percentages for some of these are shown in **Table 1** and/or are available on the Human Disease Gene website series. Additionally, the gastrointestinal symptom profile for individuals with this syndrome has been expanded to include eosinophilic esophagitis, cyclic vomiting syndrome, Mallory Weiss tears, abdominal migraine, esophageal dilation, and subglottic stenosis ^41^. A recent analysis including nine G-tube or GJ-tube fed probands demonstrated that G/GJ-tubes are overall efficacious with respect to improvements in weight gain and caregiving^41^. A recommendation has been made that OS individuals not tracking above the failure to thrive (FTT) range past 1 year of age should promptly undergo G-tube placement to avoid prolonged growth failure. If G-tubes are not immediately helping to increase weight gain after insertion, recommendations include altering formula, increasing caloric input, or exchanging a G-tube for a GJ-tube by means of a minimally invasive procedure.

### Growth, including height, weight and head circumference

Many of the OS individuals and a few of the individuals with *NAA15* variants have low height and weight, as detailed in a recent study ^41^. As demonstrated in that study, poor growth could not entirely be explained by inadequate caloric intake, inability to properly chew or swallow, growth hormone deficiency, or low appetite; past the age of 6-12 months, calorie tracking, caloric supplementation, and using feeding therapy to successfully teach individuals how to chew, swallow, and no longer choke on food did not induce adequate weight gain. Low appetite was also not an observable cause of poor growth, as multiple proband parents claimed that their individuals had great appetites, were constantly hungry, and that “desire to eat has never been a problem.” Growth hormone deficiency was treated with growth hormone (GH) administration in two probands but these efforts kept these probands in the failure to thrive (FTT) range, despite some slight weight improvements. Effect of GH administration on weight gain among GH-deficient OS individuals with growth failure should be further investigated in subsequent studies.

### Endocrine and Metabolic features

Some of these features are shown in **Table 1**. Extensive laboratory panels assessing digestive function, screening for metabolic decompensation, and hormonal dysregulation have not been significantly useful in improving the care or outcome for these individuals. Although some parents report that their children are quite lean with thermoregulation abnormalities, there is no metabolic or quantitative data yet to prove this claim. Although evidence from mouse models^43^ indicates dysregulation of glucose and insulin levels, the one human proband (Individual 46) tested thus far with extensive glucose monitoring over 14 days did not show any major abnormalities. Mouse models also show decreased brown adipose tissue, which might offer an explanation for the thermoregulation issues ^43^, but brown adipose tissue amounts have not yet been assessed in affected individuals, particularly as there are logistical challenges with getting children and adults with severe intellectual disability to cooperate with imaging exams and it is very difficult to justify the use of sedation in a research setting. It is therefore up to each individual treating clinician to decide whether to pursue clinical testing to explore whether these individuals are indeed lean and/or have decreased brown adipose tissue.

There are occasional findings in particular individuals that may or may not be related to the variants in *NAA10* or *NAA15*. For example, Individual 8 with an *NAA15* variant has been found to have corneal calcifications and persistent serum hypercalcemia, although there is no other laboratory-based evidence for hyperparathyroidism.

### Ophthalmologic and visual features

The majority of *NAA10* individuals present with visual abnormalities, which often include astigmatism, myopia, strabismus, and/or amblyopia (also see **Table 1**). Further, there are a subset of individuals who have exotropia/esotropia, microphthalmia, are blind, and/or have abnormalities to the optic disc or nerve. In *NAA15* individuals, visual abnormalities were present in a small subset as strabismus, amblyopia, astigmatism, and other visual impairment. Many individuals with this syndrome wear eyeglasses and due to this high prevalence of visual impairment, eye examination by ophthalmology is warranted. Further, thirteen individuals with *NAA10* variants and one male with p.Asn864Ser in NAA15 presented with cortical or cerebral visual impairment (CVI). CVI can be described as a “verifiable visual dysfunction which cannot be attributed to disorders of the anterior visual pathways or any potentially co-occurring ocular impairment” ^44^. CVI affects the processing of visual information, which has a profound impact on their ability to learn and must be thoroughly assessed to cater to the unique needs of each child and their impairment. Generally, most individuals have learning disabilities, and particularly seem to have difficulty with depth perception and discerning contrast, which may contribute to their difficulty using stairs or walking off of curbs. CVI was not thoroughly investigated for this paper and should be further explored with more examinations of these individuals to understand how their CVI impacts their learning and/or visuomotor abilities.

Individual 23 is a female with the common p.Arg83Cys missense in *NAA10*, yet she is the only female published to date who was born with microphthalmia, with a very small right eyeball along with a smaller, misshapen pupil, leading to the need for a prosthetic eyeball (see **Supplementary Figure 2**). She also has stigmatism in her left eye, along with myopia and possible cortical visual impairment. Her exome sequencing did not reveal any other possible cause for the microphthalmia. Individual 54 (a male) with p.Thr152Argfs*6 in *NAA10* also has microphthalmia (see **Supplementary Figure 3**), similarly to a previously published case ^39^, whereas the male in Japan did not have microphthalmia ^44^. Individual 55 has p.Glu181Alafs*67 and does not have microphthalmia, indicating that even frameshift variants toward the C-terminus of NAA10 can have quite variable outcomes.

After the posting of an earlier version of this manuscript as a preprint ^45^, one clinician (A.C.) reported to us the discovery of one female (19 years old, Individual 20 in Supplementary Information) with unilateral microphthalmia with a *NAA15* variant (NM_057175.4:c.852G>A p.(Trp284*), heterozygous). This is further proof that the expression of many phenotypes is likely titrated by the overall amounts of monomeric NAA10 and/or NatA activity, whereby microphthalmia occurs in both syndromes when some threshold is crossed during development in which likely some substrate is not acetylated enough.

### Neurologic features

The prototypical subject displays intellectual disability, global developmental delays, gross and/or fine motor delay, hypotonia, and speech delay. In many cases speech delay was severe enough for speech to be considered absent. Additionally, the presence of tonic-clonic seizures was not uncommon. Virtually all individuals have neonatal hypotonia, and several had neurogenic scoliosis. For Individual 45, intervention for neurogenic scoliosis was attempted with placement of rods. The percentages for some of the neurologic features are shown in **Table 1** and/or are listed on the Human Disease Gene website series.

Neuroimaging has been obtained in a number of these individuals, primarily to screen for brain anomalies associated with their hypotonia and growth failure, and there is a paper in preparation regarding detailed analysis of brain MRI findings. The majority possessed at least one brain abnormality with corpus callosum hypoplasia and microcephaly being by far the most common findings followed by periventricular leukomalacia (see **Supplementary Table 1** for exact figures). In some cases, there was a finding of cerebral atrophy. However, there was no radiographic evidence to explain their hypotonia or growth failure. It seems warranted to obtain brain MRI with diffusion tensor imaging, to check for white matter delays, in the first few years of life, and then a follow-up MRI around puberty. Given that some of the individuals develop seizures, it seems that baseline EEG should be obtained to check for any seizure activity and, ideally, an annual EEG, or at least one every 2-3 years, should be performed up until their early 20’s. Prophylactic treatment with anti-epileptic medication should be considered if EEG shows any possible seizures. Lastly, given that some of the individuals have developed autism, it is recommended to perform standard autism screening assessments during childhood, which can include Autism Diagnostic Inventory (ADI) or Autism Diagnostic Observation Schedule (ADOS-2) screening.

### Cognitive and psychiatric features

The psychiatric profile of subjects with OS typically includes harmful behavior which is either outwardly directed (agitation or violent behavior toward others) or inwardly directed / self-injurious (hair pulling, self-biting, head-banging, etc) in addition to impulsive or compulsive behavior. The presence of any combination of short attention span, poor eye contact, and autistic, stereotyped or repetitive behaviors (tics, hand-wringing, hand-flapping, etc) were common. The percentages for some of these features are shown in **Table 1** and/or are available on the Human Disease Gene website series.

Vineland-3 functional assessment revealed substantial variability among the *NAA10* and *NAA15* probands, although the overall level of functioning for the *NAA15* probands was significantly higher than the *NAA10* probands (**Figure 3A**, **Supplementary Table 5**). This is consistent with the results presented previously in **Figure 2** demonstrating that more *NAA10* probands were given a “severe” intellectual disability diagnosis. When the results for NAA10 were separated by sex, inheritance pattern, and variant type, it became clear that the frameshift variants located toward the C-terminal end of NAA10 have much less impact on overall functioning. One of the males (Individual 7) with a maternally inherited missense variant (c.215T>C, p.Ile72Thr) in NAA10 had an almost normal level of functioning, with an ABC standard score of 94, which is less than one standard deviation away from the “normal” of 100. However, Individual 8 (now deceased) had the exact same variant, but his ABC standard score was 59, demonstrating again that there is substantial variability even among the exact same variants. For the females with the p.Arg83Cys missense in NAA10, there is also substantial phenotypic variability with functioning as well, which appears to be equivalent in scope to the many other variants in females (**Figure 3B**). As such, there are clearly other factors affecting the overall trajectory of these individuals, in addition to just the one variant they have in *NAA10* or *NAA15*, which as mentioned above could include, among other things, the cardiac arrhythmias, feeding difficulties, delayed speech, delayed motor development, seizures, and cortical visual impairment. Comprehensive analyses of the many different Vineland sub-scale, age-equivalent scores (AES), and growth-scale values (GSVs) ^46^ falls outside the scope of the current paper and will be reported in a future paper.

**Figure 3.**
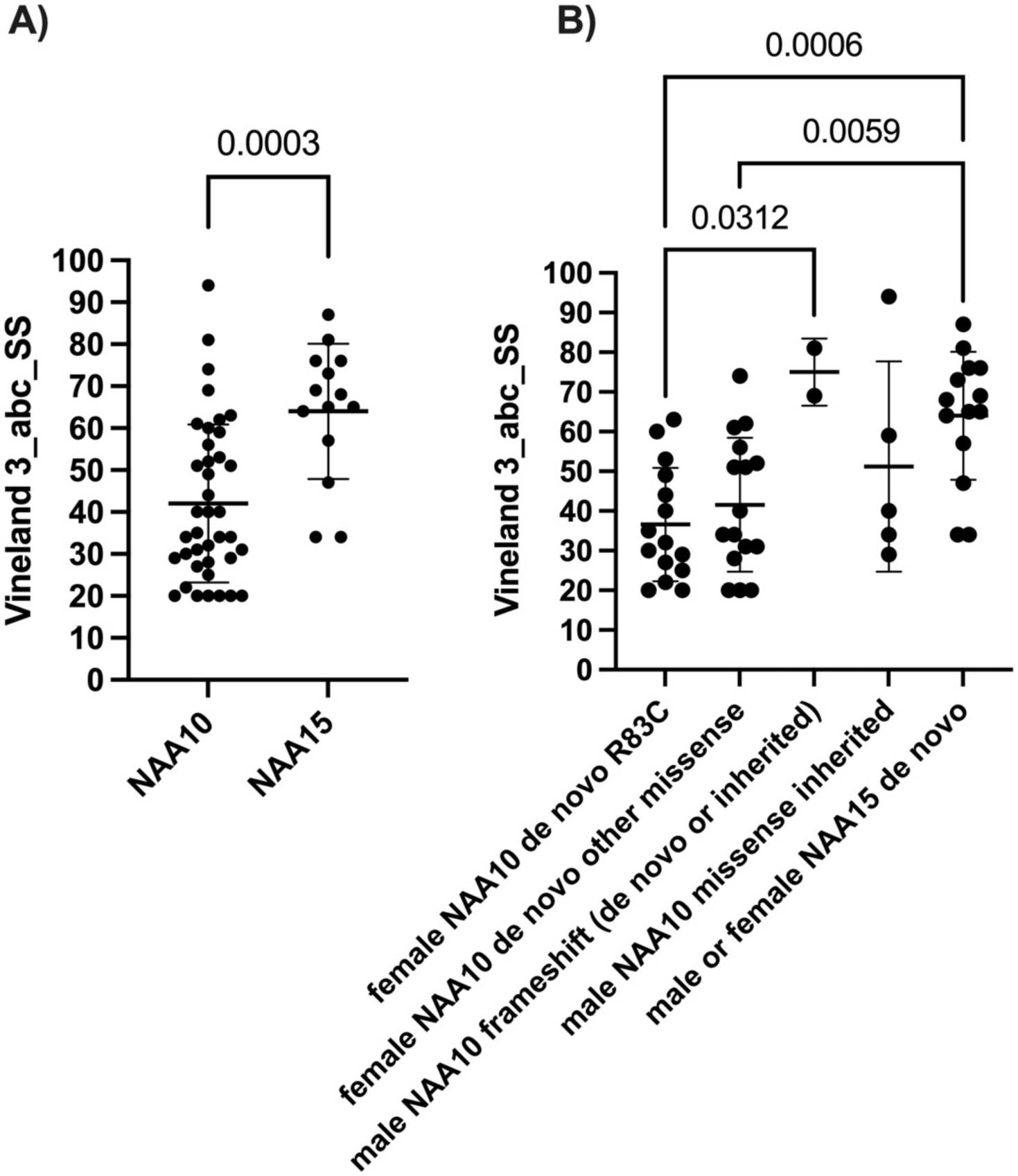
Vineland results. Adaptive Behavior Composite (ABC) Standard scores have a mean of 100 in the “normal” population and SD of 15. **A)** All scores are plotted for *NAA10* and *NAA15*, without regard for sex, inheritance pattern, or type of variant. **B)** These same scores are now separated out by sex, inheritance pattern and type of variant. In the case of *NAA15*, which is an autosomal gene, the sexes are still grouped together.

### Human Phenotype Ontology-based clustering analyses

Hierarchical clustering analyses were performed using the HPO terms downloaded from the Human Disease Gene website series in October 2022 (see **Supplementary Table 1**). The two syndromes for *NAA10* and *NAA15* do cluster separately, albeit with some overlap (see **Supplementary Figure 6**). Most importantly, and consistent with the clinical impression of these two syndromes, there was no obvious sub-clustering among the different variants within each syndrome. This provides quantitative and supportive data regarding the phenotypic spectrum for the presentation for variants in *NAA10* and *NAA15*, and this is also consistent with the presentation of NAA15-related neurodevelopmental syndrome being somewhat distinct (and usually much more mild) than NAA10-related neurodevelopmental syndrome.

### Molecular Analysis of the Variants

Most *NAA10* variants identified to date in *NAA10*-related neurodevelopmental diseases reduce NAA10- and NatA-type activity ^20^. Although novel variants clinically described here have not been characterized biochemically, it is possible to anticipate biochemical implications resulting from variants in the conserved region of NAA10 (residues 1-160) based on previous studies^13, 16, 18–21, 23, 32, 33^. However, variants and truncations occurring in the C-terminus of NAA10 require additional studies to understand the role of the NAA10 C-terminal domain. The observation that frameshifts occurring at the C-terminal region of NAA10 are associated with a lower impact on cognitive function is consistent with the conserved NAA10 domain being the minimal region necessary for *in vitro* activity.

To contextualize the phenotypes associated with the NAA10 and NAA15 mutant proteins, we inspected two published human NatA crystal structures: the heterodimeric complex composed of the NAA10 and NAA15 subunits (PDB: 6C9M) and the heterotrimeric complex composed of NAA10, NAA15, and the regulatory subunit, HYPK (PDB: 6C95). Based on these structures, the *NAA10* variants p.Pro8Ser/Asp10Glu, p.Tyr31Cys, p.His120Pro, p.Ser123Pro, p.Phe128Ser, and p.Arg149Trp would likely compromise the folding and/or thermal stability of NAA10 **(Figure 4)** ^5^. Specifically, the mutations to proline (p.His120Pro, p.Ser123Pro) occur in helices, likely destabilizing the helical structure, and the p.Tyr31Cys and p.Phe128Ser mutations likely disrupt hydrophobic core interactions that are mediated by Tyr31 and Phe128. The double variant, [c.22C>T;30C>G] p.[Pro8Ser;Asp10Glu] in the exon/intron boundary, leads to a reduction in NAA10 formation by altering the splicing of the mRNA from this allele (**Supplementary Figure 1)**.

**Figure 4.**
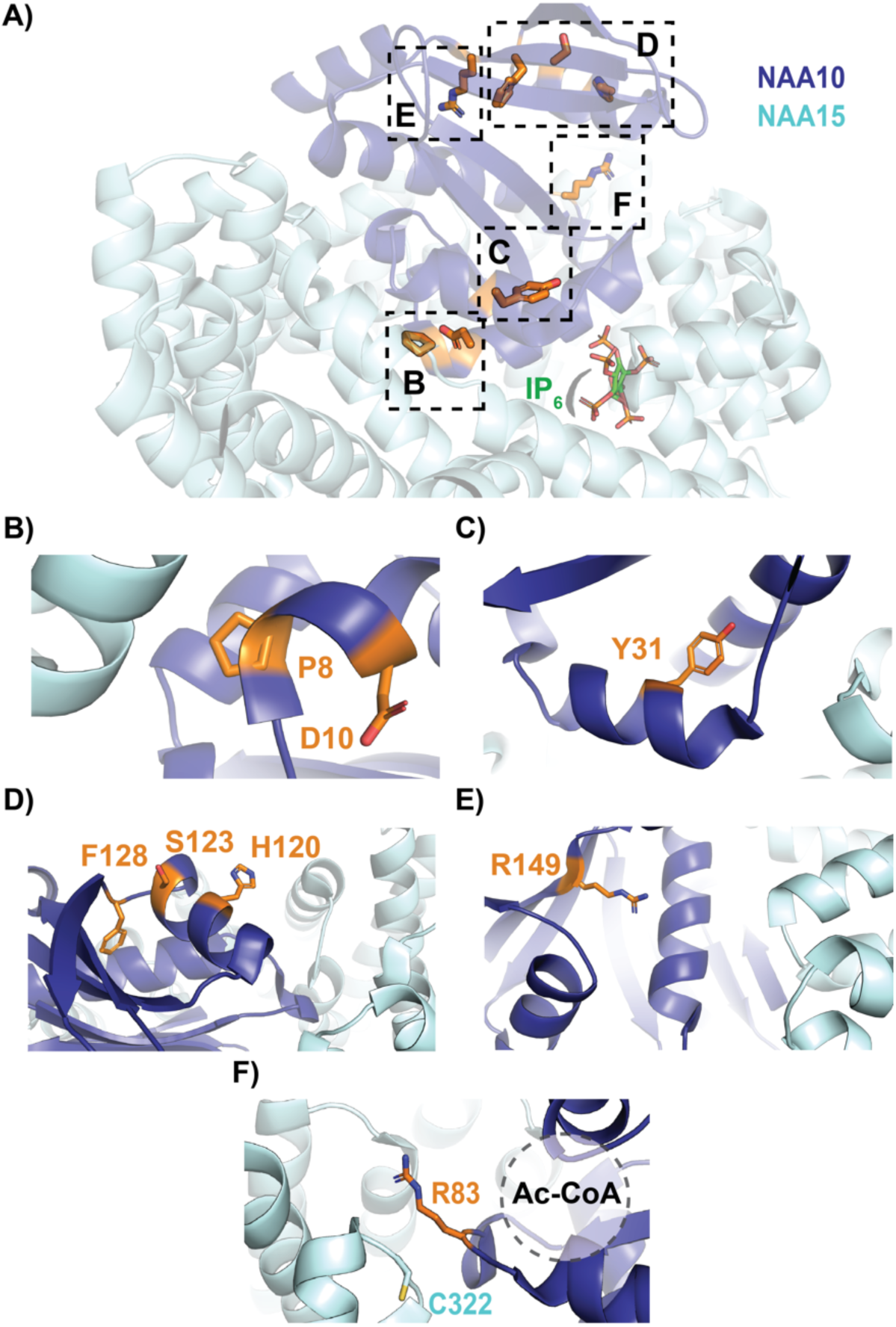
Crystal structure of human NaA complex labeled with relevant NAA10 residues. NAA10 (dark blue) and NAA15 (light cyan) are shown in cartoon with wild-type residues in stick (orange). **A)** Top view of complex annotated with relevant NAA10 residues outlined by dashed boxes: **B)** P8 and D10; **C)** Y31; **D)** H120, S123, and F128; **E)** R149; and **F)** R83 and NAA15 C322 (light cyan, stick), where the acetyl CoA binding pocket is indicated by a dash-outlined circle.

The biochemistry of the p.Arg83Cys variant is unique amongst the NAA10-type variants. The monomeric NAA10 p.Arg83Cys protein is less active than wild type NAA10 ^33^ and the heterodimeric p.Arg83Cys NatA complex was found have enhanced activity, while HYPK binding returns p.Arg83Cys NatA activity back to wild type levels ^20^. In the context of the heterodimeric NatA complex, the binding of acetyl CoA to p.Arg83Cys NAA10 is compromised, as indicated by the loss of acetyl CoA-mediated stabilization ^20^. For the wild type NAA10, the positively charged p.Arg83Cys side chain interacts with the negatively charged acetyl CoA pantothenic arm, which would be absent with the Cys variant. In addition, it is possible that this complex NatA phenotype arises from the formation of a disulfide bond between NAA10 p.Arg83Cys with the nearby NAA15 p.Cys322 residue. This disulfide bond has been proposed to anchor the complex so that the complex is better positioned to catalyze the N-terminal acetylation reaction. This disulfide bond would not form in the monomeric NAA10 state, rendering mutant NAA10 p.Arg83Cys less active than the wild type monomeric NAA10.

In contrast to the reported *NAA10* variants, *NAA15* variants tend to confer a range of biochemical and biophysical effects on NatA function ^20^. Similar to *NAA10* variants, several novel variants have been included that have not yet been biochemically characterized (p.Arg27Gly, p.Ala585Thr, p.Ala719Thr, and p.Asn864Ser). Variants occurring in NAA15 could affect NAA15 folding and/or thermostability, as well as intermolecular interactions with NAA10 (directly or through a bridging inositol hexaphosphate, IP6, molecule) or HYPK – and, thereby, NatA catalytic activity, or NatA localization to the ribosome.

Based on structural considerations of the human NatA and NatA/HYPK crystal structures **(Figure 5)** and previous *in vitro* studies, ^46^ we anticipate that the NAA15 p.Lys450Glu mutant would disrupt the NAA10-IP6 interaction and, thus, destabilize the NAA10/NAA15 complex, while the p.Lys338Asn mutant could disrupt NAA15 thermostability by disrupting a hydrogen bond between two adjacent helices in NAA15. p.Ala585Thr is disordered in both crystal structures and mutants p.Arg27Gly and p.Asp112Asn are solvent-exposed on NAA15, so it is unclear how these mutations impact NatA function. However, it is possible that they disrupt the NatA-ribosome interaction. In particular, p.Arg27 and p.Ala585 are both located in regions previously implicated in ribosome-binding, thereby altering NatA co-translational activity ^47^.

**Figure 5.**
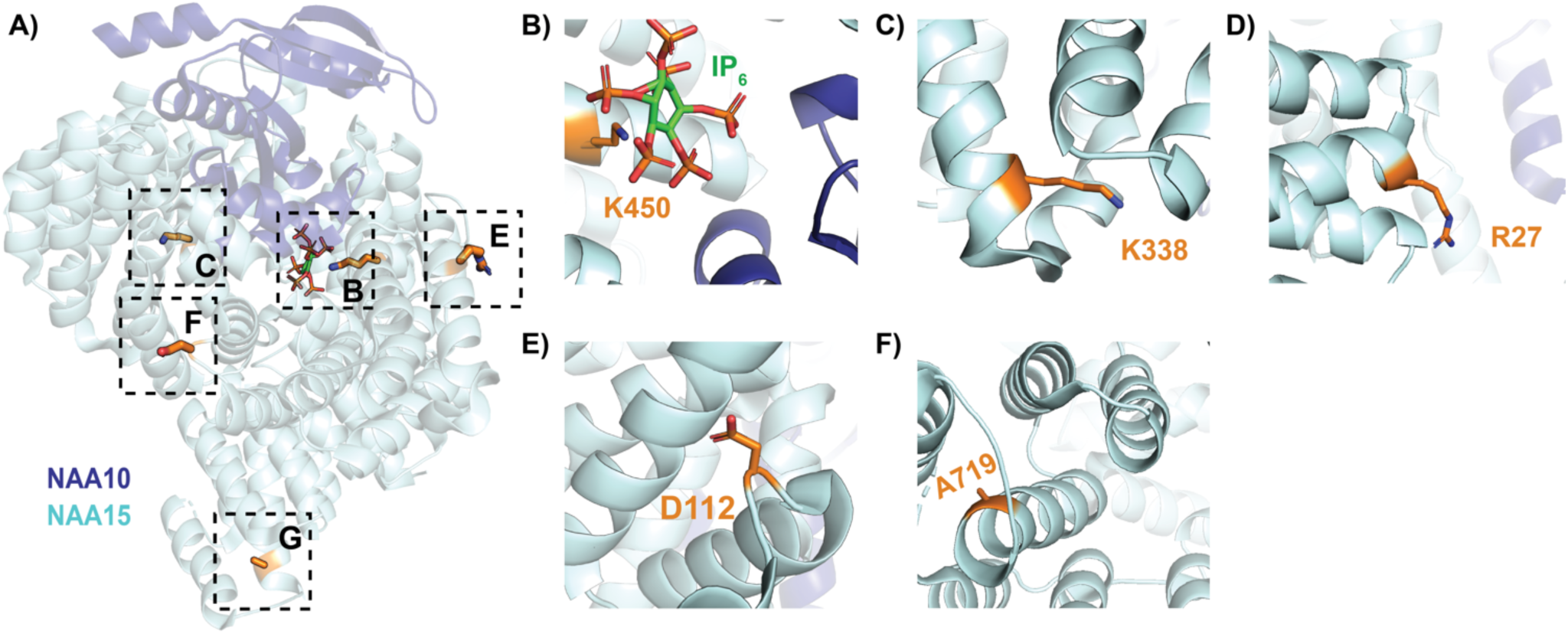
Crystal structure of human NatA complex labeled with relevant NAA15 residues. NAA10 (dark blue) and NAA15 (light cyan) are shown in cartoon with wild-type residues in stick (orange). **A)** Side view of complex annotated with relevant NAA15 residues outlined by dashed boxes: **B)** K450 with bound inositol hexaphosphate (IP6, stick); **C)** K338N; **D)** R27; **E)** D112; and **F)** A719.

P.Ala719Thr is buried within a bundle of helices in the metazoan-conserved NAA15 C-terminus, which is critical for NatA heterodimer stabilization and HYPK regulatory subunit binding.^47^ Finally, the molecular basis for the p.Asn864Ser mutant is unclear because published NatA structures are not resolved past residue 841. However, there are two published phosphorylation sites p.Ser855 and p.Ser856 in this region that have not been evaluated but may be impacted by this variant.

## Discussion

Mendelian diseases are anything but simple, as the presentations can be quite complex and variable due to modification by genetic background and environmental influences ^48^. This is quite apparent in the case of *NAA10*-related neurodevelopmental syndrome, as the variability is enormous, with different trajectories that are impacted by whether the child overcomes certain hurdles, such as learning to feed themselves and to walk. Males with *NAA10*-related neurodevelopmental syndrome may have symptoms at different ages: those with arrhythmias or other serious cardiac conditions may be embryonic lethal; others are apparent at birth experiencing cardiac concerns, hypotonia and dysmorphic features; and others come to attention later with relatively nonspecific developmental/intellectual and growth impairments.

Likewise, some females can present with similar issues as males, including rarely microphthalmia and/or prolonged QT interval, and the majority of the females with *NAA10* variants have severe to profound intellectual disability, whereas the individuals with *NAA15* variants are much less severely impaired (see **Figure 3**).

An area for future investigation concerns the amount of physical, speech, occupational, and other therapies that should be recommended. Based on these extensive interviews with the families, the anecdotal information seems to suggest that individuals with *NAA10*-related neurodevelopmental syndrome benefit from the following therapies on at least a weekly basis in at least the first 7 years of life, so as to ensure that they can eat, walk, and ideally verbalize: Mobility therapy; speech and language therapy; sensory integration therapy; hydrotherapy; occupational therapy with a focus on fine motor skills; and cortical visual impairment therapy (if warranted). The trajectory of these individuals seems to vastly improve with intensive therapy, whereas some of these individuals do not make adequate progress without adequate therapy. As mentioned, future studies can investigate this question much more quantitatively with a natural history study, documenting the exact modalities and frequencies of therapies.

One limitation of our study is that only the first nine research participants were interviewed in-person and physically examined by medical doctors, including by a medical geneticist, a neurologist, a psychiatrist, and an optometrist. After this time, all interviews were converted to videoconferencing starting in March 2020, with the onset of the COVID19 pandemic, and the families met with the psychiatrist only (GJL). However, we did not observe any major differences in the information obtained from the in-person visits as compared to the videoconference visits, and all visits included review of available medical records. Please see the **Supplementary Discussion** for additional details.

To date, most NAA10 and NAA15 variants have been studied *in vitro* using either immunopurified overexpressed full-length NAA10 (235 residues) protein from yeast ^16^ or mammalian culture ^19, 21, 23^, *S. frugiperda* (*Sf*9)-expressed recombinant NatA complex containing a C-terminally truncated NAA10 (1-160) ^20^, or an affinity-tagged full-length NAA10 expressed in *E. coli* ^48^. Each of these methodologies have their limitations. Protein isolated from mammalian cell culture offers a unique advantage, where, depending on the immunoprecipitation conditions, NatA likely co-purifies with relevant binding partners, such as the NatA regulatory protein, HYPK, and the NatE catalytic subunit, NAA50. However, this does not allow for selective isolation of the NAA10 catalytic subunit or the NatA complex without either of its binding partners. By contrast, *Sf*9 overexpression of the human NatA complex can be performed with and without the co-transfection of the HYPK virus to allow for selective production of the NatA and NatA/HYPK complexes. In addition, it is also possible to prepare purified NatE complex with or without HYPK^49^. However, this approach has its limitations because the Sf9-expressed NatA complex construct contains a C-terminally truncated NAA10 (1-160 of 235 total residues). This C-terminal region can be phosphorylated at 6 sites ^50^, and could potentially influence the activity and the stability of NAA10, although the interplay of these phosphorylation events with NAA10 and NAA15 variants has not yet been evaluated. While recombinant NAA10 overexpression in *E. coli* allows for the studying of the full-length monomeric NAA10, proteins produced in *E. coli* are not post-translationally modified. Overall, these *in vitro* assays are limited in scope as there could very well be cellular and organismal phenotypes that result from decreased (or otherwise altered) amino-terminal acetylation of a wide range of substrates.

NatA plays a complex role in the cell with a range of functional roles and varying substrate types, depending on the enzyme’s oligomeric state. This is particularly apparent with the NAA10 p.Arg83Cys variant. The heterodimeric p.Arg83Cys NatA complex has enhanced activity, HYPK binding returns p.Arg83Cys NatA activity back to wild type levels ^20, 25, 49^ while the monomeric mutant NAA10 features a diminished level of activity^20, 25, 50^. Despite this, the phenotype of the individuals with this particular mutation is not appreciably different from the other individuals reported herein. There are multiple ophthalmologic abnormalities in these females, and individual 23 is a female with the p.Arg83Cys missense in NAA10, with microphthalmia, just like Individual 54 (a male) with p.Thr152Argfs*6 in *NAA10* also with microphthalmia, and similar in phenotype to previously published cases ^20, 25, 51^. The cognitive functioning of the females with p.Arg83Cys missense in NAA10 is also similarly impaired as the other females with different missense changes (**Figure 3**), and there is a phenotypic spectrum including multiple organ systems in all of these females with missense changes in NAA10.

These pathologies highlight the need to build off the groundwork laid by focused biochemical studies into model systems that can account for NatA’s multiple oligomeric states and cellular functions. One such multidimensional approach has been to study the human mutations in the human genes expressed in a *S. cerevisiae* model knocked out for endogenous yeast *Naa10* and/or *Naa15* ^50^. However, this approach has its own caveats. For example, *S. cerevisiae* is a much simpler unicellular organism that does not represent the complexity of a human cell or a human being, including the absence of expression of the NatA regulatory subunit, HYPK. Instead, these mutations are perhaps best studied using patient-derived or cell lines with the mutations engineered into them, as has been done recently with *NAA15* ^51^, and/or with animal models with the human variants engineered into *NAA10* or *NAA15*.

In conclusion, we have presented phenotypic information on many new variants in *NAA10* and *NAA15*, along with additional phenotypic information on some previously published *NAA10* variants. We document an extensive phenotypic spectrum caused by likely decreased expression and/or function of NAA10 and/or the NatA complex, with the cases involving *NAA10* variants usually presenting as more severe than cases involving *NAA15*. Although some papers have also suggested that there might be different allelic presentations or mechanisms of action for different variants involving *NAA10*, such as with microphthalmia present in males with splice-site ^51^ or frameshift ^53^ variants, the present study demonstrates that this is much more likely to be a phenotypic spectrum of one unitary disease. Although some do refer to this entire disease entity as Ogden syndrome, another name could be *NAA10*-related neurodevelopmental syndrome. The latter name is longer and thus more cumbersome, whereas the name Ogden syndrome is more memorable and easier to introduce when describing the disease to non-experts, but the longer name does follow the recently suggested dyadic nomenclature ^54^.

Further information about this can be found in **Supplementary Discussion**. We further suggest that the disease entity involving *NAA15* variants should be labeled as *NAA15*-related neurodevelopmental syndrome.

## Data Availability

All data produced in the present study are available upon reasonable request to the authors. The sequencing data were generated as part of clinical testing, so the underlying raw data are not consented for deposition to a public database.

https://humandiseasegenes.nl/naa10/

https://humandiseasegenes.nl/naa15/

https://db.gestaltmatcher.org/

## Acknowledgements

The authors would like to thank the Genome Aggregation Database (gnomAD) and the groups that provided exome and genome variant data to this resource. A full list of contributing groups can be found at http://gnomad.broadinstitute.org/about. Early in the project, Drs. Milen Velinov, Ricardo Madrid, and Roseanne Ricciardi graciously performed medical genetic, neurology, and optometry examinations, respectively, on the nine families with *NAA10* variants seen in-person at the Jervis Clinic. GJL would also like to thank the Clinical and Administrative staff of the Jervis Clinic for their assistance, and the many clinicians who performed tests and/or wrote medical records that were obtained and summarized. This study has made use of data generated by Human Disease Genes Website series, www.humandiseasegenes.nl.

## Author Contributions

GJL was responsible for all videoconferencing and primary data collection, with secondary summaries performed by EM, MV, TB, AP, KS, IB, and MT. Data analysis was performed by GJL MV, TB, and AP. Bioinformatics analyses were performed by QL and KW. In-person examinations at the Jervis Clinic were aided by MG and KA. Biomedical interpretation of the human NatA structure were provided by LG and RM. Vineland-3 psychological testing was performed by EHI. The first draft of the manuscript was written by GJL, MV, TB, and AP, with input thereafter from all other authors.

## Ethical Approval

Both oral and written patient consent were obtained for research and publication, with approval of protocol #7659 for the Jervis Clinic by the New York State Psychiatric Institute - Columbia University Department of Psychiatry Institutional Review Board. Written family consent was given for publication of any photography of the children.

## References

1. Arnesen, T. et al. Proteomics analyses reveal the evolutionary conservation and divergence of N-terminal acetyltransferases from yeast and humans. Proc. Natl. Acad. Sci. U. S. A. 106, 8157–8162 (2009).

2. Starheim, K. K., Gevaert, K. & Arnesen, T. Protein N-terminal acetyltransferases: when the start matters. Trends Biochem. Sci. 37, 152–161 (2012).

3. Arnesen, T. et al. Identification and characterization of the human ARD1-NATH protein acetyltransferase complex. Biochem. J. 386, 433–443 (2005).

4. Arnesen, T. et al. The chaperone-like protein HYPK acts together with NatA in cotranslational N-terminal acetylation and prevention of Huntingtin aggregation. Mol. Cell. Biol. 30, 1898–1909 (2010).

5. Gottlieb, L. & Marmorstein, R. Structure of Human NatA and Its Regulation by the Huntingtin Interacting Protein HYPK. Structure 26, 925–935.e8 (2018).

6. Arnesen, T. et al. Characterization of hARD2, a processed hARD1 gene duplicate, encoding a human protein N-alpha-acetyltransferase. BMC Biochem. 7, 13 (2006).

7. Gendron, R. L. et al. Loss of tubedown expression as a contributing factor in the development of age-related retinopathy. Invest. Ophthalmol. Vis. Sci. 51, 5267–5277 (2010).

8. Dörfel, M. J. & Lyon, G. J. The biological functions of Naa10 - from amino-terminal acetylation to human disease. Gene 567, 103–131 (2015).

9. Aksnes, H., Ree, R. & Arnesen, T. Co-translational, Post-translational, and Non-catalytic Roles of N-Terminal Acetyltransferases. Mol. Cell 73, 1097–1114 (2019).

10. Kweon, H. Y. et al. Naa12 compensates for Naa10 in mice in the amino-terminal acetylation pathway. Elife 10, (2021).

11. Blomen, V. A. et al. Gene essentiality and synthetic lethality in haploid human cells. Science 350, 1092–1096 (2015).

12. Wang, T. et al. Identification and characterization of essential genes in the human genome. Science 350, 1096–1101 (2015).

13. Rope, A. F. et al. Using VAAST to identify an X-linked disorder resulting in lethality in male infants due to N-terminal acetyltransferase deficiency. Am. J. Hum. Genet. 89, 28–43 (2011).

14. Lyon, G. J. Personal account of the discovery of a new disease using next-generation sequencing. Interview by Natalie Harrison. Pharmacogenomics 12, 1519–1523 (2011).

15. Gogoll, L. et al. Confirmation of Ogden syndrome as an X-linked recessive fatal disorder due to a recurrent NAA10 variant and review of the literature. Am. J. Med. Genet. A 185, 2546–2560 (2021).

16. Van Damme, P., Støve, S. I., Glomnes, N., Gevaert, K. & Arnesen, T. A Saccharomyces cerevisiae model reveals in vivo functional impairment of the Ogden syndrome N-terminal acetyltransferase NAA10 Ser37Pro mutant. Mol. Cell. Proteomics 13, 2031–2041 (2014).

17. Dörfel, M. J. et al. Proteomic and genomic characterization of a yeast model for Ogden syndrome. Yeast 34, 19–37 (2017).

18. Myklebust, L. M. et al. Biochemical and cellular analysis of Ogden syndrome reveals downstream Nt-acetylation defects. Hum. Mol. Genet. 24, 1956–1976 (2015).

19. Bader, I. et al. Severe syndromic ID and skewed X-inactivation in a girl with NAA10 dysfunction and a novel heterozygous de novo NAA10 p.(His16Pro) variant - a case report. BMC Med. Genet. 21, 153 (2020).

20. Cheng, H. et al. Phenotypic and biochemical analysis of an international cohort of individuals with variants in NAA10 and NAA15. Hum. Mol. Genet. (2019) doi:10.1093/hmg/ddz111.

21. McTiernan, N. et al. NAA10 p.(N101K) disrupts N-terminal acetyltransferase complex NatA and is associated with developmental delay and hemihypertrophy. Eur. J. Hum. Genet. 29, 280–288 (2021).

22. Ree, R. et al. A novel NAA10 p.(R83H) variant with impaired acetyltransferase activity identified in two boys with ID and microcephaly. BMC Med. Genet. 20, 101 (2019).

23. Støve, S. I. et al. A novel NAA10 variant with impaired acetyltransferase activity causes developmental delay, intellectual disability, and hypertrophic cardiomyopathy. Eur. J. Hum. Genet. 26, 1294–1305 (2018).

24. Afrin, A., et al. NAA10 variant in 38-week-gestation male patient: a case study. Cold Spring Harb Mol Case Stud 6, (2020).

25. Johnston, J. J. et al. NAA10 polyadenylation signal variants cause syndromic microphthalmia. J. Med. Genet. jmedgenet-2018 (2019).

26. Gupta, A. S., Saif, H. A., Lent, J. M. & Couser, N. L. Ocular Manifestations of the NAA10- Related Syndrome. Case Rep. Genet. 2019, 8492965 (2019).

27. Maini, I. et al. Clinical Manifestations in a Girl with NAA10-Related Syndrome and Genotype–Phenotype Correlation in Females. Genes 12, 900 (2021).

28. Cheng, H. et al. Truncating Variants in NAA15 Are Associated with Variable Levels of Intellectual Disability, Autism Spectrum Disorder, and Congenital Anomalies. Am. J. Hum. Genet. 102, 985–994 (2018).

29. Ward, T. et al. Mechanisms of congenital heart disease caused by NAA15 haploinsufficiency. Circ. Res. 128, 1156–1169 (2021).

30. Dingemans, A. J. M. et al. Human disease genes website series: An international, open and dynamic library for up-to-date clinical information. Am. J. Med. Genet. A 185, 1039–1046 (2021).

31. Richards, S. et al. Standards and guidelines for the interpretation of sequence variants: a joint consensus recommendation of the American College of Medical Genetics and Genomics and the Association for Molecular Pathology. Genet. Med. 17, 405–424 (2015).

32. McTiernan, N. et al. Biochemical analysis of novel NAA10 variants suggests distinct pathogenic mechanisms involving impaired protein N-terminal acetylation. Hum. Genet. (2022) doi:10.1007/s00439-021-02427-4.

33. Saunier, C. et al. Expanding the Phenotype Associated with NAA10-Related N-Terminal Acetylation Deficiency. Hum. Mutat. 37, 755–764 (2016).

34. Resnik, P. Using information content to evaluate semantic similarity in a taxonomy. arXiv [cmp-lg] (1995).

35. Couto, F. M. & Lamurias, A. Semantic Similarity Definition. in Encyclopedia of Bioinformatics and Computational Biology 870–876 (Elsevier, 2019).

36. Hsieh, T.-C. et al. GestaltMatcher facilitates rare disease matching using facial phenotype descriptors. Nat. Genet. (2022) doi:10.1038/s41588-021-01010-x.

37. Hustinx, A. et al. Improving Deep Facial Phenotyping for Ultra-rare Disorder Verification Using Model Ensembles. in Proceedings of the IEEE/CVF Winter Conference on Applications of Computer Vision 5018–5028 (2023).

38. Casey, J. P. et al. NAA10 mutation causing a novel intellectual disability syndrome with Long QT due to N-terminal acetyltransferase impairment. Sci. Rep. 5, 16022 (2015).

39. Shishido, A. et al. A Japanese boy with NAA10-related syndrome and hypertrophic cardiomyopathy. Hum. Genome Var. 7, 23 (2020).

40. Chaudhary, P., Ha, E., Vo, T. T. L. & Seo, J. H. Diverse roles of arrest defective 1 in cancer development. Arch. Pharm. Res. 42, 1040–1051 (2019).

41. Sandomirsky, K., Marchi, E., Gavin, M., Amble, K. & Lyon, G. J. Phenotypic variability and Gastrointestinal Manifestations/Interventions for growth in Ogden syndrome (also known as NAA10-related Syndrome). medRxiv (2022) doi:10.1101/2022.03.16.22272517.

42. 42. Gmail, L. @. & Hinton, G. Visualizing Data using t-SNE. https://www.jmlr.org/papers/volume9/vandermaaten08a/vandermaaten08a.pdf?fbcl (2008).

43. Lee, C.-C. et al. Naa10p inhibits beige adipocyte-mediated thermogenesis through N-α- acetylation of Pgc1α. Mol. Cell 76, 500–515.e8 (2019).

44. Chokron, S., Kovarski, K. & Dutton, G. N. Cortical Visual Impairments and Learning Disabilities. Front. Hum. Neurosci. 15, 713316 (2021).

45. Lyon, G. J. et al. Expanding the Phenotypic spectrum of Ogden syndrome (NAA10-related neurodevelopmental syndrome) and NAA15-related neurodevelopmental syndrome. medRxiv (2022) doi:10.1101/2022.08.22.22279061.

46. Farmer, C. A. et al. Person Ability Scores as an Alternative to Norm-Referenced Scores as Outcome Measures in Studies of Neurodevelopmental Disorders. Am. J. Intellect. Dev. Disabil. 125, 475–480 (2020).

47. Magin, R. S., Deng, S., Zhang, H., Cooperman, B. & Marmorstein, R. Probing the interaction between NatA and the ribosome for co-translational protein acetylation. PLoS One 12, e0186278 (2017).

48. 48. Lyon, G. J. & O’Rawe, J. Human genetics and clinical aspects of neurodevelopmental disorders. in The Genetics of Neurodevelopmental Disorders. (ed. Mitchell, K.) (Wiley), 2015).

49. Deng, S., McTiernan, N., Wei, X., Arnesen, T. & Marmorstein, R. Molecular basis for N- terminal acetylation by human NatE and its modulation by HYPK. Nat. Commun. 11, 818 (2020).

50. Olsen, J. V. et al. Quantitative phosphoproteomics reveals widespread full phosphorylation site occupancy during mitosis. Sci. Signal. 3, ra3 (2010).

51. Esmailpour, T. et al. A splice donor mutation in NAA10 results in the dysregulation of the retinoic acid signalling pathway and causes Lenz microphthalmia syndrome. J. Med. Genet. 51, 185–196 (2014).

52. Wu, Y. & Lyon, G. J. NAA10-related syndrome. Exp. Mol. Med. 50, 85 (2018).

53. Sidhu, M., Brady, L., Tarnopolsky, M. & Ronen, G. M. Clinical Manifestations Associated With the N-Terminal-Acetyltransferase NAA10 Gene Mutation in a Girl: Ogden Syndrome. Pediatr. Neurol. 76, 82–85 (2017).

54. Biesecker, L. G. et al. A dyadic approach to the delineation of diagnostic entities in clinical genomics. Am. J. Hum. Genet. 108, 8–15 (2021).

